# Validation of L-Type Calcium Channel Blocker Amlodipine as a Novel ADHD Treatment through Cross-Species Analysis, Drug-Target Mendelian Randomization, and clinical evidence from medical records

**DOI:** 10.1101/2024.05.30.24308216

**Authors:** Haraldur Þorsteinsson, Hannes A. Baukmann, Hildur S. Sveinsdóttir, Dagmar Þ. Halldórsdóttir, Bartosz Grzymala, Courtney Hillman, Jude Rolfe-Tarrant, Matthew O. Parker, Justin L. Cope, Charles N. J. Ravarani, Marco F. Schmidt, Karl Æ. Karlsson

## Abstract

ADHD is a chronic neurodevelopmental disorder which significantly affects life outcomes. First-line treatments carry the risk of adverse side effects and present a high abuse potential, coupled with a 25% rate of non-response, necessitating novel treatments. Here, we validate amlodipine as an ADHD treatment using model rats and zebrafish and human genetic data. Amlodipine reduced hyperactivity in the Open Field Test in SHR rats and reduced both hyperactivity and impulsivity in the 5-Choice Serial Reaction Time Task in *adgrl3.1^-/-^*zebrafish. We show that amlodipine also passes the blood brain barrier and reduces telencephalic activation. Mendelian Randomization analysis using human genetic data revealed significant associations between ADHD and genetic variations in the subunits of L-type calcium channels (α1-C; CACNA1C, β1; CACNB1, α2δ3; CACNA2D3), and the combined genes targeted by amlodipine. Finally, we show that amlodipine mitigates key ADHD symptoms in a cohort of people with a high ADHD genetic liability. Given its well-tolerated profile, its efficacy in mitigating both hyperactivity and impulsivity across different species, coupled with genetic evidence from human data, the potential utility of amlodipine as a novel treatment for human ADHD is compelling.

## Introduction

Attention-deficit/hyperactivity disorder (ADHD) is a prevalent neurodevelopmental condition impacting 2-5% of the global population.^1^ Symptoms typically emerge in early childhood and persist into adulthood, leading to a range of behavioral and psychiatric challenges.^2^ Left untreated, ADHD is associated with functional impairments, heightened risk of mood and anxiety disorders, and severe consequences, including an increased risk of suicide.^3^ The primary treatment for ADHD involves stimulant medications like methylphenidate and d-amphetamines. However, these medications come with side effects such as appetite loss, hypertension, headaches, and sleep disturbances, as well as high abuse liability.^4^ Second-line options also carry adverse effects, provide less efficacy than stimulants, and around 25% of patients show no significant improvement from any therapeutic.^5,6^ Thus, there is an urgent need for more innovative and effective ADHD therapeutics.

We previously identified five compounds with the potential to be repurposed for ADHD treatment: aceclofenac, amlodipine, doxazosin, moxonidine, and LNP599.^7^ Here, our aim was to investigate the potential for these previously identified compounds to be adopted as viable ADHD therapeutics. Towards this aim, first, we determined the efficacy in a mammalian ADHD model, using an Open Field Test (OFT) in the Spontaneously Hypertensive Rat (SHR).^8^ Second, we determined if the compounds are effective for rescuing impulsivity (not only hyperactivity) using the 5-Choice Serial Reaction Time Task (5-CSRTT) in adult mutant zebrafish.^9,10^ We show effects of other L-type calcium channel blockers (LTCCBs), determine brain penetration and effects on c-Fos expression. Finally, we determine human translational value using Mendelian randomization (MR)^11,12^ and by assessing the effect of amlodipine on mental health questionnaire responses associated with high ADHD polygenic risk scores (PRS).

## Results

### Amlodipine affects Open Field Behavior in SHR rats

To assess the therapeutic effects of amlodipine on ADHD, we examined its effects on the gold standard rodent ADHD assay. Eight groups were used: seven SHR groups, each group receiving a different treatment: vehicle control, clonidine (positive control), moxonidine, LNP599, doxazosin, amlodipine, and aceclofenac. A control group of Wistar-Kyoto (WKY) rats received the vehicle control only. Our data show that long-term daily administration (day 0 to day 29) of amlodipine induces significant behavioral changes in SHR rats in the OFT; the effects of all other drugs were non-significant (Supplementary Figure 1) and thus excluded from further analysis. Specifically, amlodipine administration reduced indices of hyperactivity, i.e., distance traveled and ambulatory time (Figure 1A-I). No significant differences in rearing time were observed between vehicle-treated and amlodipine-treated female or male SHR rats were observed on either day 0 or day 29 (n.s.) (Figure 1A-C). For distance traveled, a significant interaction between time and treatment was evident among female rats (*F*(2,26) = 10.08, *p* < 0.001) and when sexes were combined (*F*(2,56) = 5.222, *p* < 0.01). No significant interaction was observed among male rats (*F*(2,27) = 0.2071, *p* = 0.8142). At day 29, female amlodipine-treated SHR rats showed reduced distance traveled compared to vehicle-treated SHR female rats (*p* = 0.0021) (Figure 1D). For ambulatory time, a significant difference between vehicle-treated SHR and amlodipine-treated SHR rats (*p* < 0.01) also appeared when pooling the sexes (Figure 1F). A significant interaction between time and treatment was evident among female rats (*F*(2, 26) = 17.47, *p* < 0.0001) and when sexes were combined (*F*(2, 56) = 9.127, *p* < 0.001). No significant interaction was observed among male rats (*F*(2, 27) = 0.3082, *p* = 0.7373). At day 29, female amlodipine-treated SHR rats showed reduced ambulatory time compared to vehicle-treated SHR rats (*p* < 0.01) (Figure 1G). Among male rats, significant differences were evident at day 29 between vehicle-treated SHR (*p* < 0.0001) and amlodipine-treated SHR rats (*p* < 0.0001) (Figure 1H). Similarly, when combining males and females, a significant effect was evident at day 29 between vehicle-treated SHR and amlodipine-treated SHR rats (p < 0.01) (Figure 1I).

**Figure 1.**
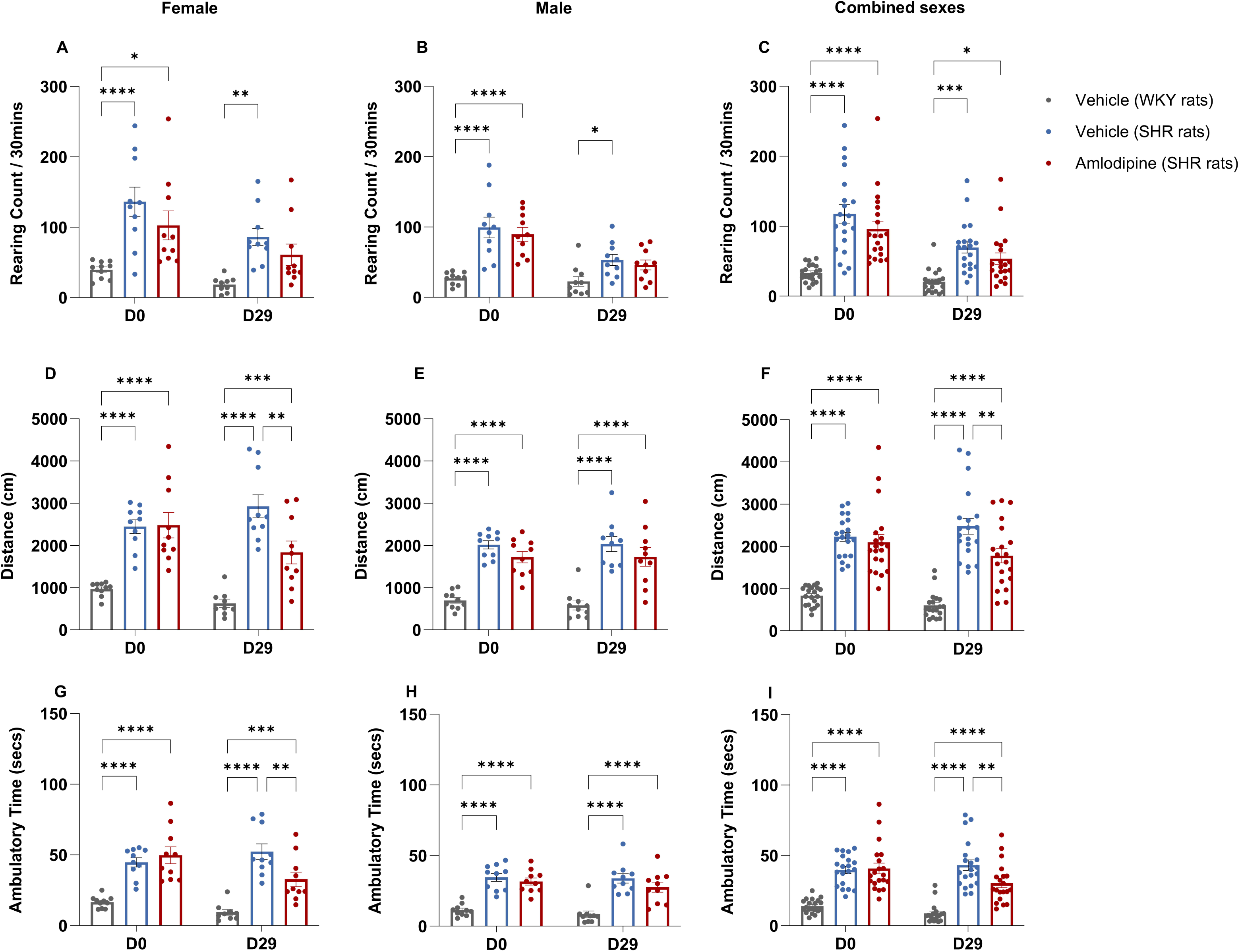
Effect of amlodipine on Open Field Behavior in SHR rats. SHR rats were treated with either amlodipine (10 mg/kg) or a vehicle daily for 30 days; WKY rats received vehicle only. Assessments were made before treatment (D0) and at end of treatment (D29) using OFT. Measurements included rearing frequency (A, B, C), distance traveled in the center zone (D, E, F), and ambulatory time in the center zone (G, H, I). Reduced distance traveled was revealed at D29, with amlodipine-treated SHR rats covering significantly less distance than vehicle-treated SHR rats in the female group (D) and for both sexes combined (F). Additionally, reduced ambulatory time was observed at D29, where amlodipine-treated SHR rats spent significantly less time ambulating in the center zone compared to vehicle-treated SHR rats in the female group (G) and for both sexes combined (I). Data are depicted as mean ± SEM, \**P* ≤ 0.05, \*\**P* ≤ 0.01, \*\*\**P* ≤ 0.001, \*\*\*\**P* ≤ 0.0001.

In summary, there is an effect of amlodipine on distance moved and ambulatory time but not rearing frequency in the OFT. This effect is driven by female rats. Since only amlodipine was effective in modulating the behavior of SHR rats in the OFT, only amlodipine was used in subsequent experiments.

### Amlodipine reduces impulsivity in adult *adgrl3.1^-/-^* zebrafish

We have previously shown that zebrafish lacking the adhesion G-protein coupled receptor latrophilin 3 (*adgrl3.1^-/-^*) show increased hyperactivity in the open field and increased impulsivity in the 5-CSRTT.^10^ Like in SHR rats, we previously found that amlodipine effectively rescues hyperactive phenotype in *adgrl3.1^-/-^* zebrafish^7^ models of ADHD. Here, we tested the hypothesis that amlodipine would also reduce impulsivity in ADHD zebrafish models in the 5-CSRTT - an assay specifically designed to gauge impulsivity^13^ and pharmacologically validated in *adgrl3.1^-/-^* ADHD models.^10^ There were no differences in Phase 2 training between wild-type (WT) and *adgrl3.1^-/-^* (main effect Strain LMM: *F*[1,297.06] = 0.09, *p* = 0.769). During baseline 5-CSRTT Phase 2 (pre-drug treatment), *adgrl3.1^-/-^*fish showed more premature responses than WT, *t*(40.45) = 2.02, *p* = 0.05, 95%CI = 0 - 0.1 (Figure 2B). Drug treatment was initiated following Phase 2. There was a significant main effect of Drug Type (LMM: *F* [2,16.55] = 3.76, *p* = 0.045), with treatment with both amlodipine (*p* < 0.001) and methylphenidate (*p* < 0.01) causing a reduction in impulsive responses in the *adgrl3.1^-/-^* fish (Figure 2C).

**Figure 2.**
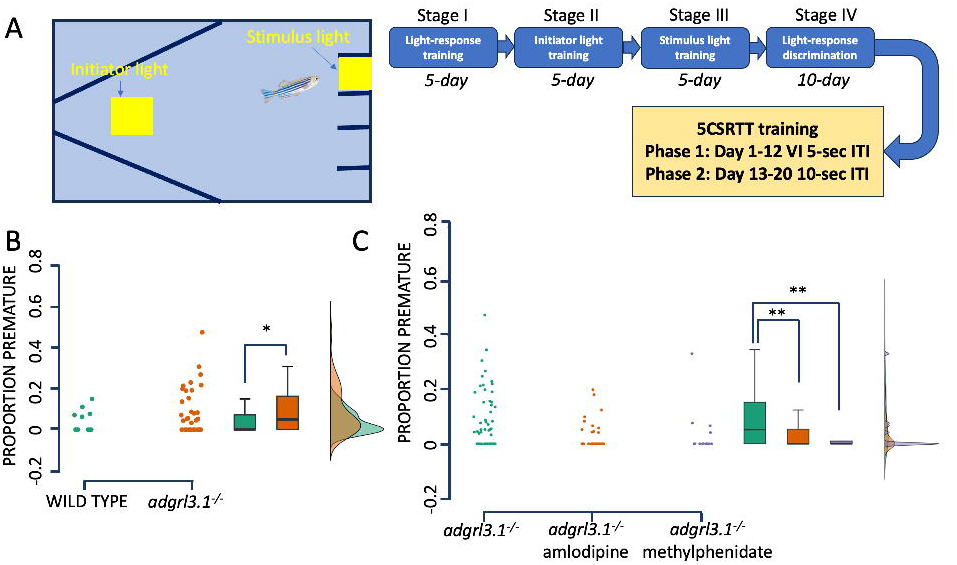
Acute effects of amlodipine and methylphenidate on impulsivity in adult zebrafish. (A) The test tank and schematic overview of the 5-CSRTT. (B) Proportion of premature responses in age matched wild-type and *adgrl3.1^-/-^*adult zebrafish. (C) Effects of amlodipine (10 µM) and methylphenidate (10 µM) on impulsive responses in *adgrl3.1^-/-^*adult zebrafish. Data are depicted as mean ± SEM.,\**P* = 0.05; \*\**P* < 0.01.

### Effects of additional L-type calcium channel blockers on hyperactivity and sleep in larval *agdrl3.1^-/-^* ADHD model

Due to the efficacy of amlodipine in rescuing hyperactivity and impulsivity in ADHD rodent and zebrafish models, twelve other LTCCBs were assayed in the larval ADHD assay described previously.^7^ Five LTCCBs (diltiazem, lacidipine, nicardipine, nisoldipine and nitrendipine) did not have any effect on hyperactivity whereas seven (cilnidipine, felodipine, isradipine, nifedipine, nilvadipine, nimodipine and verapamil) did. The effective LTCCBs generated a profile roughly similar to that of amlodipine: that is reduction of hyperactivity with low effects on sleep parameters. However, all drugs except nifedipine and nimodipine were toxic at medium to high doses and induced higher behavioral variability than amlodipine (Supplementary Figure 2).

### Amlodipine crosses the blood-brain barrier and affects c-Fos expression

To assess the extent to which amlodipine can cross the blood-brain barrier (BBB), and to establish the functional mechanisms by which amlodipine may exert its behavioral effects in the central nervous system, we quantified the concentration of amlodipine in the brain tissue and examined its impact on neuronal activation in the telencephalon. Following exposure to 10 µM amlodipine (n = 6), whole-brain amlodipine levels in adult *adgrl3.1^-/-^*fish were measured at 0.013 ± 0.003 ng/ml; no amlodipine was detected in non-exposed fish (except for one which measured at 9 x 10^-6^ ng/ml, which we attribute to sample contamination) (n = 6). No sex differences were measured. One-way ANOVA revealed significant differences in c-Fos expression within the telencephalon (*F*(3,88) = 16.76, *p* < 0.0001). *adgrl3.1^-/-^* and WT larvae subjected to amlodipine treatment showed significant downregulation of c-Fos expression compared to those treated with DMSO (*p* < 0.05 and *p* < 0.01, respectively). These results show, at the dose used to rescue the hyperactive and impulsive phenotype, amlodipine permeates the brain and suppresses c-Fos expression (Figure 3).

**Figure 3.**
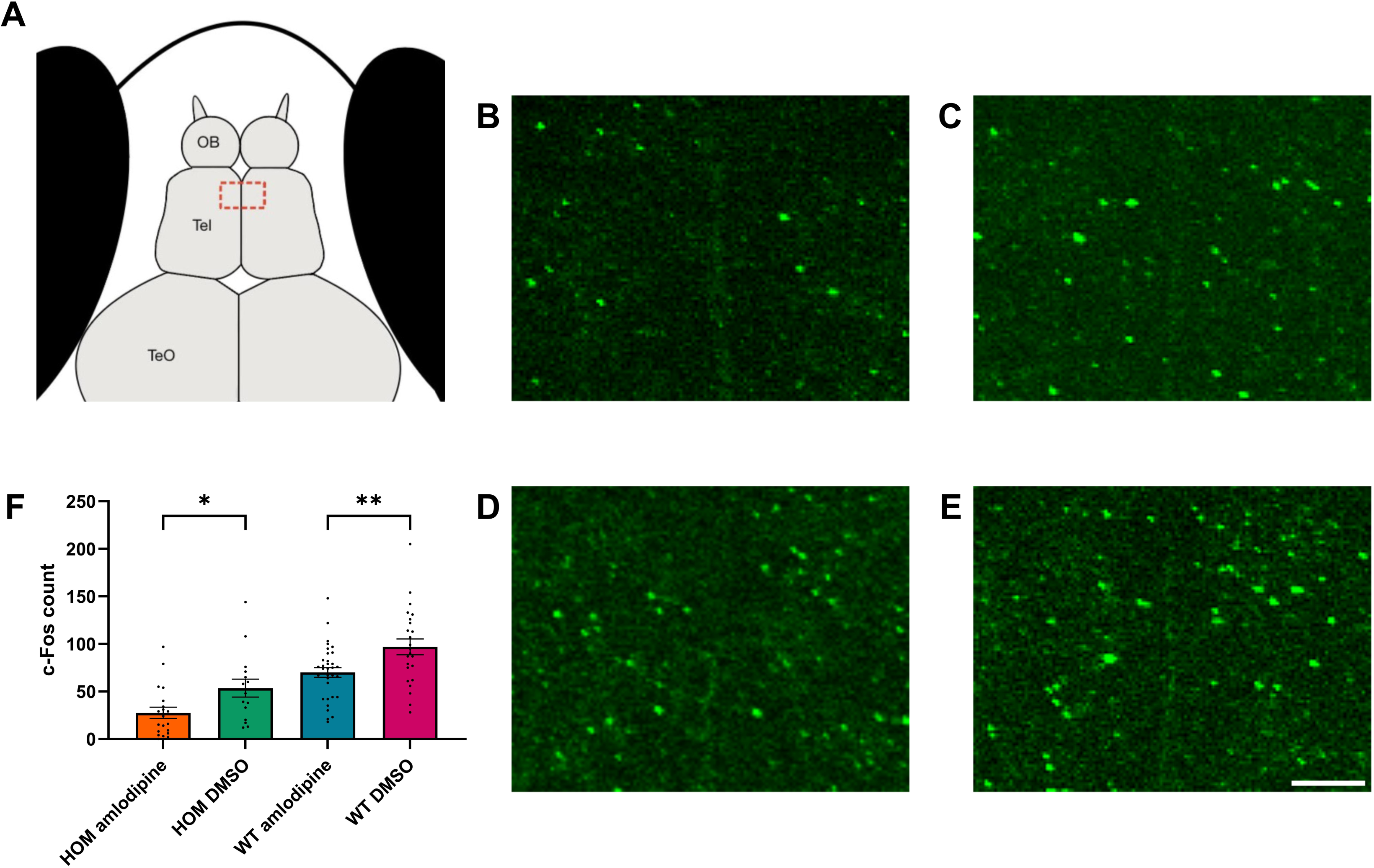
c-Fos expression in *adgrl3.1^-/-^* zebrafish in the telencephalon following amlodipine exposure. (A) A schematic representation of a 6 dpf zebrafish larva, red dashed rectangle within the telencephalon denotes the area used for counting. Representative figures of immunohistochemically labeled c-Fos of *adgrl3.1^-/-^* zebrafish larvae treated with (B) 10 µM amlodipine and (C) 0.1% DMSO and WT zebrafish larvae treated with (D) 10 µM amlodipine and (E) 0.1% DMSO. (F) c-Fos count for each group. Data are mean ± SEM, \**P* ≤ 0.05, \*\**P* ≤ 0.01, (scale bar = 10 µm).

### Genome-wide Mendelian randomization on hypertension and ADHD

Having established the effects of amlodipine on ADHD symptoms across two animal models, and confirmed its ability to cross the BBB and its putative functional effects in the telencephalon, we next employed MR to extend the results from animal studies to human genetic data. We first confirmed the well-established effect of amlodipine on hypertension before exploring the relationship between amlodipine target perturbation and ADHD. First, the causal effect of 140 traits on hypertension and ADHD was estimated using genome-wide MR. The manually curated list of traits includes blood cell counts, metabolites, amino acids, lipids, and enzymes. Three traits with a significant causal effect estimate were found for hypertension (Figure 4A, Supplementary Table 2): BMI and phenylalanine increased the risk of hypertension, while nucleated erythrocyte percentage decreased it. Three traits with a significant causal effect estimate for ADHD (Figure 4B, Supplementary Table 2) were identified: sodium in the urine, higher BMI and acetate increased the risk of general ADHD.

**Figure 4.**
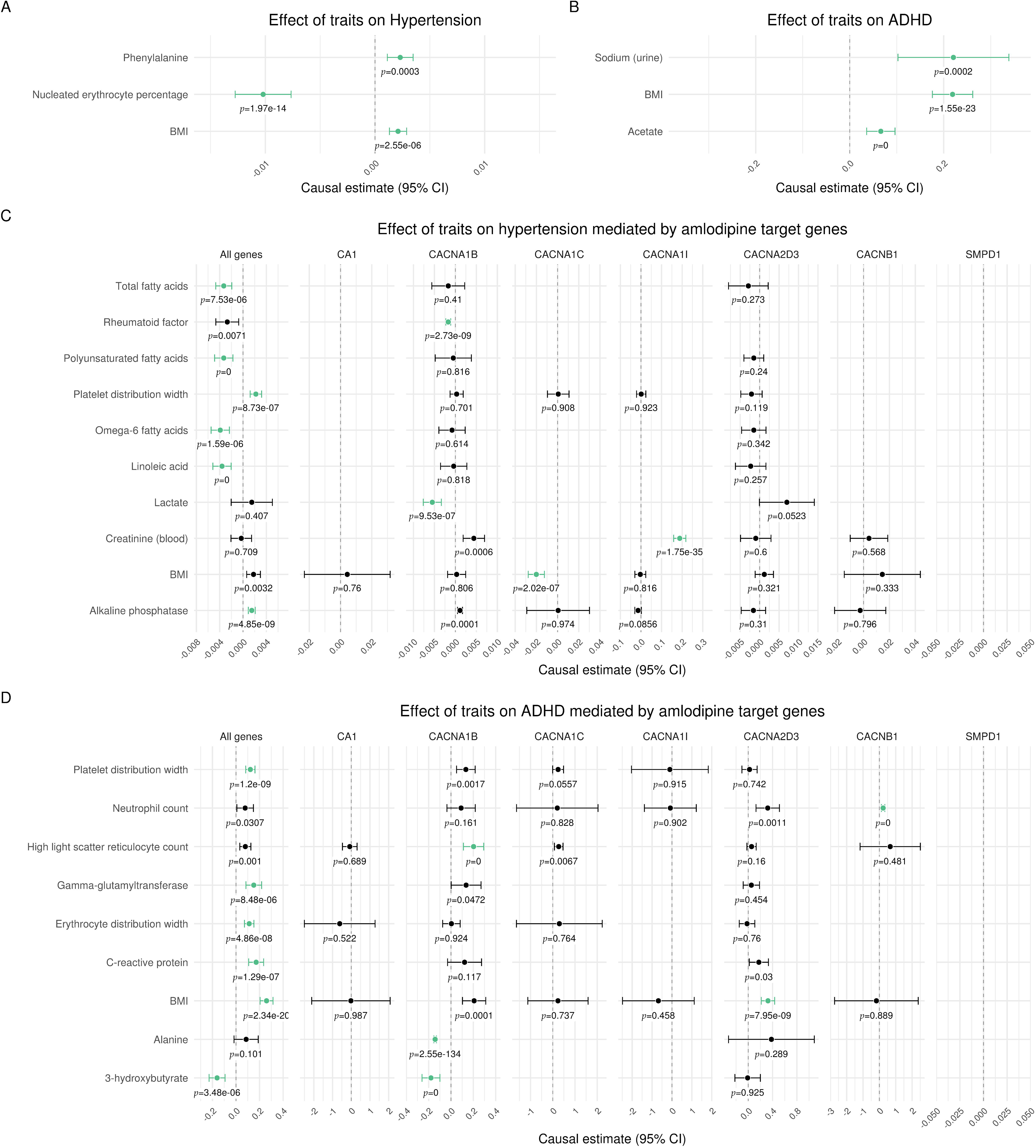
Genome-wide and drug-target Mendelian Randomization. (A) Only exposures with significant causal effects for hypertension are shown. The significance threshold was Bonferroni corrected for multiple testing of 140 traits (*p* < α/n = 0.05/140). Green dots indicate a significant causal effect without pleiotropy. Estimates are scaled per 1-standard deviation (SD) increase in the respective exposure trait, i. e., every 1-SD higher genetically proxied trait level was associated with the respective change in log odds ratio for ADHD. (B) Only exposures with significant causal effect estimates for ADHD are shown. The significance threshold was Bonferroni corrected for multiple testing of 140 traits (*p* < α/n = 0.05/140). Green dots indicate a significant causal effect without pleiotropy. Estimates are scaled per 1-standard deviation (SD) increase in the respective exposure trait, i. e., every 1-SD higher genetically proxied trait level was associated with the respective increase in log odds ratio for hypertension. (C) Only exposures which had significant causal effects for hypertension mediated by one of the eight groups of genes are reported. The significance threshold was Bonferroni corrected for multiple testing of 140 traits and 8 gene selections (p < α/n = 0.05/140/8). Green dots indicate a significant causal effect without pleiotropy. Estimates are scaled per 1-standard deviation (SD) increase in the respective exposure trait, i. e., every 1-SD higher genetically proxied trait level was associated with the respective decrease in log odds ratio for hypertension. (D) Only exposures which had significant causal effects for ADHD mediated by one of the eight groups of genes are reported. The significance threshold was Bonferroni corrected for multiple testing of 140 traits and 8 gene selections (p < α/n = 0.05/140/8). Green dots indicate a significant causal effect without pleiotropy. Estimates are scaled per 1-standard deviation (SD) increase in the respective exposure trait, i. e., every 1-SD higher genetically proxied trait level was associated with the respective increase in log odds ratio for ADHD.

### Drug-target Mendelian randomization confirms potential for amlodipine to target ADHD symptoms

Amlodipine inhibits the voltage-dependent calcium channels of type L (subunits α_1_-C and β_1_, encoded by *CACNA1C* and *CACNB1*, respectively), type T (subunit α_1_-I, *CACNA1I*), type N (subunit α_1_-B, *CACNA1B*) and auxiliary subunit α_2_δ_3_ (*CACNA2D3*). In addition to calcium channels, amlodipine inhibits Carbonic anhydrase 1 (*CA1*) and Sphingomyelin phosphodiesterase (*SMPD1*), according to Drugbank online (https://go.drugbank.com/drugs/DB00381). To investigate the potential effect of amlodipine on ADHD, the instrumental variables for the MR experiments were drawn only from the seven genes that code for the protein targets of amlodipine (drug-target MR). Eight groups of experiments were conducted: drawing genetic variants from the genes of all amlodipine drug targets combined and the genes of the seven individual drug targets. The same selection of exposures was screened.

First, the known effect of amlodipine on hypertension was reconfirmed using our new method: significant effects on hypertension were caused by the variance in the genes encoding for the α_1_ subunits of all three calcium channel subtypes (L-type subunit α_1_-C, *CACNA1C*; T-type subunit α_1_-I, *CACNA1I*; N-type subunit α_1_-B, *CACNA1B*) and by all amlodipine target genes combined (Figure 4C, Supplementary Table 3). In all cases, these effects were mediated by a unique trait. No significant effect was observed when using gene variants from *CACNB1*, *CACNA2D3*, *CA1*, or *SMPD1*. Interestingly, significant effects on ADHD were caused by the variance in the genes encoding for the three tested subunits of L-type calcium channels (subunits α_1_-C *CACNA1C*; β_1_, *CACNB1*; α_2_δ_3_, *CACNA2D3*) and by all amlodipine target genes combined (Figure 4D, Supplementary Table 4). No significant effect was observed when using gene variants from *CACNA1B*, *CACNA1I*, *CA1*, or *SMPD1*.

### Amlodipine is associated with lower self-reported ADHD symptoms in the UK Biobank, independent of hypertension

We finally examined the effects of amlodipine in treating ADHD symptoms using the UK Biobank. ADHD is not among the health outcomes recorded in the initial UK Biobank assessment interview, although 42 individuals reported medications consistent with an ADHD diagnosis. Inpatient records identify an additional 11 individuals with the relevant ICD10 diagnosis code (F90). Given these small numbers, ADHD PRS cannot be validated directly against an ADHD diagnosis. Instead, we replicated a number of associations with ADHD risk comorbidities identified by Haan et al’s^14^ Estonian Biobank PheWAS for ADHD risk. Here, logistic regression identifies our ADHD PRS as a significant predictor of hypertension, chronic obstructive pulmonary disease, type 2 diabetes, obesity, mental and behavioral disorders due to use of alcohol, depressive episode, recurrent depressive disorder, and other anxiety disorders (Supplementary Figure 3). Among the UK Biobank assessment questions, we identified one related to mood swings and another related to risk-taking as most closely aligned with the ADHD phenotype itself, independent of the comorbidities previously identified. We find that amlodipine status is a significant predictor of both outcomes in logistic regression models with ADHD risk as a covariate – in particular, when ADHD risk is taken into account, individuals taking amlodipine are significantly less likely to report mood swings or a propensity for risk taking (Figure 5). In contrast, the antihypertensive Ramipril, an ACE inhibitor, shows no such effects (Supplementary Figure 4), highlighting the efficacy of amlodipine in mitigating symptoms of ADHD, not explained by its antihypertensive effects.

**Figure 5.**
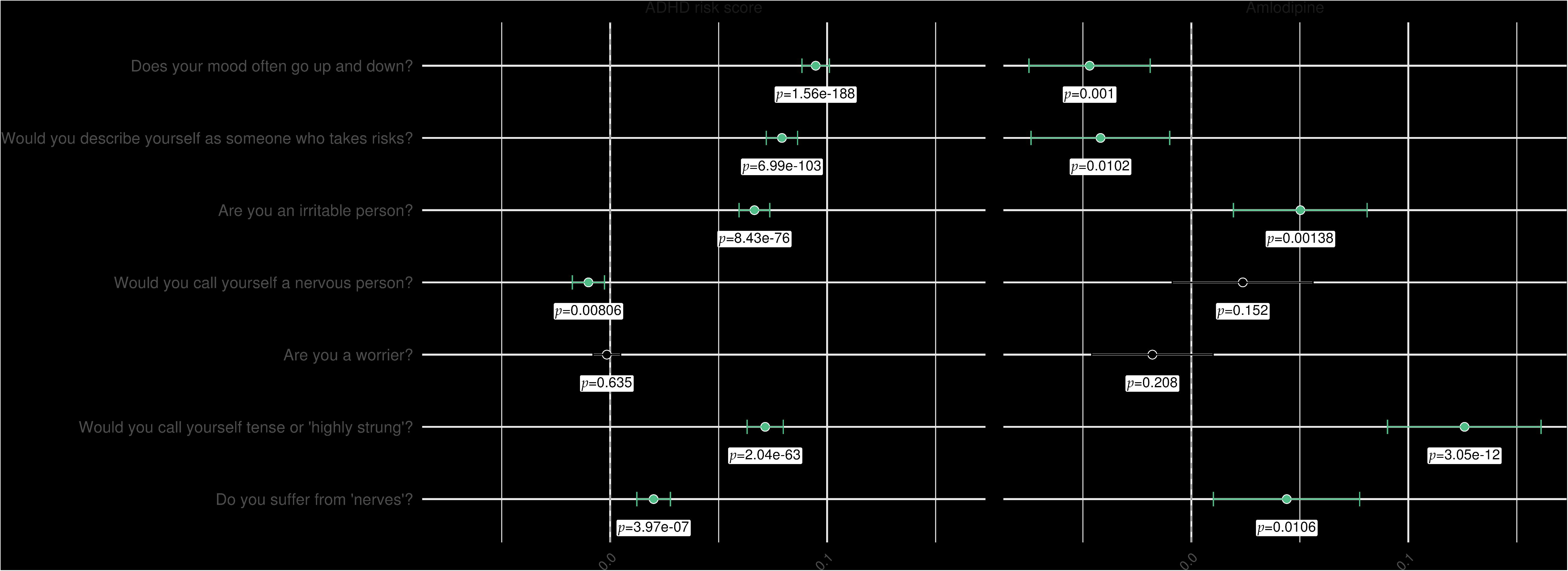
UK Biobank mental health questionnaire responses as functions of ADHD risk score and amlodipine prescription status. Each row represents a logistic regression model where UK Biobank participants’ responses to the question in the left-hand column are modeled as functions of ADHD risk score and amlodipine prescription status. The points plotted in the center and right-hand columns are the coefficient estimates for these two predictors with their corresponding p-values printed below. The bars around these points represent 95% confidence intervals for these estimates. Points and confidence interval bars in green indicate that the given predictor is significant (*p* < 0.05) in the given model.

## Discussion

In the current experiments, we used zebrafish and rodent assays to assess the pre-clinical efficacy of five previously identified novel putative ADHD therapeutics and MR to gauge human genetic support. We show that of the five candidates identified in a prior screen (aceclofenac, amlodipine, doxazosin, moxonidine and LNP599), only amlodipine demonstrated efficacy in the rodent assay. Therefore, we focused the subsequent experiments on amlodipine and, notably, used MR to demonstrate a causal correlation of variants in amlodipine target genes on ADHD in a human genetic sample.

The SHR model is widely regarded as the gold standard for studying ADHD in mammals.^8^ Therefore, we first conducted a re-assessment of the potentially therapeutic compounds previously identified using the OFT in SHR vs the WKY controls. Amlodipine did not affect rearing frequency, indicative of anxiety, but reduced two measures of hyperactivity: distance traveled and ambulatory time. Other compounds tested did not. The assay was done using a single dose so it cannot be ruled out that other compounds are efficacious using a different dose. Importantly, the fact that amlodipine was efficacious argues strongly for translatability since the assay differed greatly from the initial larval zebrafish screen: it was performed in a mammal with a different genetic background^15^ and using long-term i.p. injections, as opposed to the acute immersion administration in the zebrafish. The study thus extends the reduction of a hyperactive phenotype to a mammalian system with a different genetic background.

5-CSRTT is a behavioral assay adapted from rodents and developed to assess various aspects of attention, impulsivity, and cognitive function.^13^ It is designed to measure an animal’s ability to sustain attention, make accurate and timely responses, and inhibit impulsive responses. In the assay, impulsivity is operationalized specifically as responses occurring prior to the stimulus onset (i.e., premature or anticipatory responding). In the current experiment, this assay is used to extend the measurement of the efficacy of amlodipine from hyperactivity to impulsivity in adult zebrafish *adgrl3.1* mutants. We show that amlodipine effectively rescues impulsivity in the 5-CSRTT and the effect compares to that of methylphenidate (Cohen’s d = 0.62 and 0.59, respectively).

The efficacy of amlodipine in rescuing key features of ADHD in both rodent and zebrafish models raised the question of similar effects of other LTCCBs. We thus assayed 12 additional antihypertensive LTCCBs in the larval hyperactive screening assay.^7^ Seven of the 12 LTCCBs induced significant reduction in the hyperactivity larval zebrafish model. Neurotoxicity was common at higher doses and effects were more variable than those of amlodipine. We can conclude, first, the effects of amlodipine cannot be explained by general antihypertensive properties of LTTCBs and, second, that the effects are dependent on specific L-type calcium channel target engagement.

Amlodipine is widely reported to have a low brain penetration.^16^ Rodent data indicate approximately 20-fold lower brain penetration than nifedipine and nimodipine.^17^ The current data is hard to understand in the absence of BBB penetration. Indeed, other authors have concluded that amlodipine crosses the BBB to account for their data.^18^ Amlodipine exhibits a very slow dissociation from binding sites, suggesting that strong central effects could be achieved by brain accumulation.^19^ In the current experiment, we show that amlodipine reduces central c-Fos expression and crosses the BBB at therapeutic dose in adult *adgrl3.1*^-/-^ zebrafish. We conclude that the effects reported here are driven by the engagement of amlodipine with central L-type calcium channels.

MR is a robust and accessible tool for examining the causal relationship between an exposure and an outcome from GWAS summary statistics using genetic variants.^12^ MR leverages randomly allocated genetic variants as instrumental variables for studying the effect of varying exposure. The random allocation of genetic variants at conception means that this paradigm is less vulnerable to environmental confounding and reverse causation bias that can hinder causal inference in traditional epidemiological study designs. Naturally occurring variations in genes coding for drug target proteins can be leveraged to inform on the effect of their pharmacological perturbation.^20^ Association between a relevant genetic variant affecting the exposure and the outcome may be taken as evidence for the potential efficacy of a drug affecting the exposure pathway.^12^

We report a new approach for drug-target MR to account for polypharmacology and utilize it to examine the differential effects of amlodipine on hypertension and ADHD. We could reproduce the well-established effect of amlodipine on hypertension. Significant effects were caused by the variance in the genes encoding for the α_1_ subunits of all three calcium channel subtypes investigated as well as by all amlodipine target genes combined. This aligns with the fact that the α_1_ subunit forms the Ca^2+^ selective pore, housing both voltage-sensing machinery and the drug/toxin-binding sites. Differences in significant mediating traits suggest varying biochemical pathways. Importantly, we also found significant causal effects on ADHD, supporting our animal studies. Interestingly, these were caused only by the variance in the genes encoding for three tested subunits of L-type calcium channels and by all amlodipine target genes combined. These results suggest a major role of voltage-gated L-type calcium channels in the effect of amlodipine on ADHD.

The very low rates of ADHD diagnosis in the UK Biobank precluded the assessment of amlodipine prescription on the disorder in that cohort directly, so we constructed a PRS to estimate the genetic liability of ADHD outside of diagnosis.^14^ We cross-checked ADHD risk and associated medical conditions with amlodipine prescriptions. As reported previously, ADHD genetic liability is strongly associated with numerous medical conditions^14^ which are positively impacted by amlodipine prescriptions. Here, we found that self-reported mood swings and risk taking were reversed by amlodipine, clearly demonstrating the effect in a human population.

Amlodipine possesses several strengths for repurposing, including a well-established safety profile, low risk of drug interactions, FDA approval, affordability as a generic medication, and a known mechanism of action. Amlodipine has been reported to bind seven proteins, including subunit α_1_-C of the voltage-dependent calcium channels of type L, the coding gene of which is consistently implicated in neuropsychiatric disorders, including ADHD.^21^ LTCCs play roles in neuronal firing patterns, activity-dependent transcription regulation through calcium second messenger pathways, and the release of monoamines, affecting the prefrontal cortex^22,23^ and are, therefore, poised to modulate neural circuits important for ADHD. Amlodipine could, therefore, directly affect symptoms of ADHD via neurotransmitter release and neuronal excitability. In particular, calcium ions play a role in releasing neurotransmitters like dopamine and norepinephrine. These neurotransmitters are known to be involved in the regulation of attention, focus, and impulse control, all of which are affected in ADHD. Moreover, amlodipine is neuroprotective, beyond its blood pressure lowering effect, by modulating neuroinflammation, protecting mitochondrial structure and function and reducing oxidative stress^24–26^ and may thus affect a neural root cause of ADHD.^27,28^

In conclusion, we show that amlodipine is effective in reducing hyperactivity and impulsivity in zebrafish and rodent models of ADHD; we implemented a new approach to study polypharmacology using drug-target MR, and validated the effect of amlodipine on ADHD in human genetic data, demonstrating mitigating effects in a human population. To the best of our knowledge, this is the first drug-target MR study using instrumental variables from multiple drug target genes. We conclude that amlodipine holds a strong potential to be developed as a novel therapeutic to treat ADHD in humans.

## Methods

### SHR rat Open Field Test Housing and rodent care

WKY wild-type (n = 20) and SHR (n = 140) strains were purchased from The Center for Research Libraries, Germany. Animals ranged from 5 to 7 weeks old upon arrival and were housed in open-topped polycarbonate cages with a 13:11-h light-dark cycle under a constant temperature of 20-23 °C and relative humidity of 40-70%. All animals were provided standard chow (Teklad Global 2016) and water, each accessible ad libitum. The subjects underwent a two-week acclimation and handling period before the experimental procedures. Each subject was marked with a unique identification number using tail marks with permanent marker according to the Standard Operating Procedures of the Testing Facility. All animal experiments were performed as specified in the license authorized by the National Animal Experiment Board of Finland (ESAVI/20200/2021) and according to the National Institutes of Health (Bethesda, MD, USA) guidelines for the care and use of laboratory animals.

### Drug treatment

After the initial two-week acclimation and handling period, the SHR rats were divided into seven groups (n = 20 per group). For 30 days, these groups were administered a control vehicle (90% Acetate buffer, 10% Propylene Glycol), clonidine (0.01 mg/kg) (MedChem Express, Monmouth Junction, USA), moxonidine (0.5 mg/kg), doxazosin (8 mg/kg), amlodipine (10 mg/kg), aceclofenac (5 mg/kg) (all from Cayman Chemicals, Michigan, USA) or LNP599 (10 mg/kg) (Green Pharma, Orléans, France). A group of WKY rats (n = 20) dosed with a vehicle served as an additional control group. The administration of treatments was performed via intraperitoneal injections, delivered daily at a volume of 10 ml/kg throughout the 30 study days. Each group maintained a balanced sex distribution, with ten females and ten males.

### Open Field Tests

OFT was conducted twice for all rats: one day before initiating the dosing regimen (D0) and the 29th study day (D29). Testing occurred during the light phase between 14:00-16:00, roughly 6 hours post-dosing. Prior to the assessment, rats were given a 1-hour acclimation period in the test room. The test utilized activity chambers (Med Associates Inc, St Albans, VT; dimensions: 27 x 27 x 20.3 cm) fitted with infrared beams. Rats were individually positioned at the center of the chamber, and behavioral observations were recorded in 5-minute intervals over a 30-minute duration. Quantitative analysis was performed on the following five dependent measures: total locomotion, locomotion in the center of the open field, rearing rate in the center, total rearing frequency, and velocity. Testing conditions were optimized to minimize stress, with ambient light intensity set between 10 and 30 lux using red light. Within the enclosed chamber, light intensity ranged from 0 to 5 Lux.

### Statistical analysis

Group comparisons for distance traveled, rearing frequency, and velocity for all eight groups were analyzed using a mixed-model approach (REML), followed by a post-hoc analysis with Fisher’s LSD test without correction. Evaluations of differences in ambulatory episodes and central zone entries were performed using the same mixed-model framework (REML), followed by post-hoc analysis with Tukey’s multiple comparisons test. When evaluating the effect of amlodipine on the SHR rats in comparison to the vehicle-treated SHR and WKY rats, for distance traveled, rearing frequency, ambulatory time, and central zone entries, analyses were performed using a mixed-model approach (REML), followed by post-hoc analysis with Tukey’s multiple comparisons test.

### *adgrl3.1^-/-^*zebrafish 5-CSRTT

#### Housing and generation of fish line

Homozygous *adgrl3.1* knock-out zebrafish (*adgrl3.1^-/-^*) created through CRISPR-Cas9 (described previously^7^), alongside age-matched *adgrl3.1^+/+^* control fish were used. The fish were housed in a University of Portsmouth (UK) aquarium facility, maintained on a 14:10 light:dark cycle at 28°C (pH ∼ 8). The *adgrl3.1^-/-^*and control fish were bred concurrently through pair breeding and raised until they reached four months post-fertilization. All tanks were equipped with enrichment substrates, starting from 10 days post-fertilization (dpf). Offspring from both groups were randomly selected for experiments, maintaining equal sex distribution. All work was carried out following ethical approval from the University of Portsmouth Animal Welfare and Ethical Review Body (AWERB) and under license from the UK Home Office (PPL P9D87106F).

### Drug treatment

The *adgrl3.1^-/-^* fish received individual immersion in 10 µM amlodipine (Cambridge bioscience Ltd, Chambridge, UK) for a 30-minute duration before behavioral recording. The dose was based on the established effective dose from Sveinsdóttir et al.^7^. To administer the drug, it was dissolved in aquarium-treated water, and each fish was individually treated in 300 mL beakers. In the case of the 5-CSRTT, amlodipine treatment occurred after achieving steady-state responses in the final phase of the task.

### 5-CSRTT

Adult *adgrl3.1^-/-^* show high levels of impulsivity in the 5-CSRTT, which is rescued with atomoxetine.^10^ Here, we trained adult *adgrl3.1^-/-^*zebrafish (nD=D48; power analyses based on previous work from our group showing medium-large effect sizes on the 5-CSRTT (Figure 2 A),^10^ a validated test for measuring impulsivity in zebrafish.^13^ The task involves training the fish to respond to a light stimulus in one of five locations in a specialized testing arena (Zantiks AD, Cambridge, UK). Training on the 5-CSRTT has been reported extensively elsewhere.^13^

Briefly, initiation training comprised 4 distinct stages, each lasting ∼1-2 weeks (depending on fish performance). Fish were trained once/day in sessions that lasted 30 trials (typically ∼30-45 min). Following habituation to the test tank, in Stage 1, fish were trained that swimming over any of the lights in the tank is reinforced with a small food reward (∼2mg ZM-400 dry feed [ZM systems, Winchester, UK] delivered via an automated feeder upon correct responses). In Stage 2, fish were trained to initiate a trial by swimming over the initiator light. Again, in this stage, correct responses were reinforced with a food reward. In Stage 3, in order to be reinforced fish were required to activate one of the 5 stimulus lights, all of which are illuminated simultaneously following a correct initiator light activation. In the final pre-training stage, Stage 4, fish were required to locate and activate a single stimulus light following a correct initiator light activation. Finally, fish were trained on the 5-CSRTT over 2 phases (each lasting ∼5 days). In Phase 1, trials are initiated by activation of the initiator light, and then, following a variable-interval 5-sec inter-trial interval (ITI), one of the 5 stimulus lights is illuminated. Fish were required to swim into the correct stimulus light aperture in order to be reinforced. Commission errors, including incorrect responses (ie swimming into the incorrect aperture) or premature responses (ie swimming into any aperture prior to illumination of a stimulus light), or omission errors (ie failing to swim into an aperture during illumination of the stimulus light) resulted in no reinforcement, and a 10-sec ‘time out’. In the second stage of 5-CSRTT training, the task is identical, except the ITI is increased to VI-10-sec.

Impulsivity is assessed in the second stage of the 5-CSRTT by quantifying fish’s ability to withhold the response to the light stimulus during a defined pre-stimulus interval (usually a variable 5-second interval).

Drug treatment was carried out in a pseudo-counterbalanced order (amlodipine, methylphenidate), with a maximum of one drug exposure per week. For example, baseline recording was carried out on Monday and Tuesday, drug treatment Wednesday, followed by baseline Thursday and Friday. Drug exposures were replicated to ensure reliability. During drug treatment, fish were exposed for 30-min immediately prior to 5-CSRTT testing in a 1L tank (fish were isolated during drug treatment). They were then removed from the drug, and placed immediately into the 5-CSRTT apparatus, and training commenced.

### Statistical analysis

Data from the 5-CSRTT were analyzed in two separate analyses. First, differences between baseline responses of WT and *adgrl3.1^-/-^* fish were analyzed using a *t*-test. Second, data pertaining to drug responses (amlodipine, methylphenidate) in *adgrl3.1^-/-^* fish were analyzed using linear mixed effects models (fixed factors were drug [baseline vs. amlodipine 10 µM vs. methylphenidate 10 µM], outcomes were proportion correct responses and proportion premature responses, random effects was fish ID). Data are presented as mean ± SEM. A *p*-value threshold of less than 0.05 was set for statistical significance. All statistical analysis and graphs were generated using GraphPad software (GraphPad Software, CA, USA).

### Larval assay

*adgrl3.1^-/-^* and WT zebrafish larvae were assayed and analyzed for hyperactivity and sleep parameters as previously described.^7^ Twelve different LTCCBs were evaluated: cilnidipine, diltiazem, felodipine, isradipine, lacidipine, nicardipine, nifedipine, nilvadipine, nimodipine, nisoldipine, nitrendipine, and verapamil (Greenpharma, Orléans, France). All drugs were assessed at three different concentrations, 1 µM, 10 µM, and 30 µM, except for felodipine which was assessed at 0.1 µM, 1 µM, and 5 µM (n = 24 for each group) and compared to larvae treated with 0.3% DMSO. Each drug was recorded twice, and statistical analysis performed on the pooled data. All procedures in this study were carried out in strict compliance with the regulations of and approved by the National Bioethics Committee of Iceland (regulation 460/2017).

### Blood-brain barrier penetration

*adgrl3.1^-/-^* zebrafish (n = 12) were exposed to 10 µM of amlodipine or vehicle for a duration of 30 minutes (as above). Following the exposure, they were transferred to system water for rinsing before euthanization and brain extraction. Waters UPLC I-Class Plus (Waters Corporation, CT, USA) tandom Q Exactive high-resolution mass spectrometer (Thermo Fisher Scientific, MA, USA) was used to analyze the brain samples for quantification of amlodipine. Four ion transitions, 238.6 m/z, 294.2 m/z, 377.2 m/z, and 392.1 m/z, were considered, and the quantitative verification of amlodipine required at least two ion transitions for confirmation.

### C-Fos imaging

*adgrl3.1^-/-^* and WT zebrafish larvae (5 dpf) were exposed to 10 µM of amlodipine or 0.1% DMSO for 24 hours in a 96-well plate (n = 20 per group). Immediately following exposure, immunohistochemical staining was conducted, as detailed in Sveinsdóttir et al.^29^ Fluorescent images were acquired using an Olympus IX33 confocal microscope (Olympus, Tokyo, Japan). For c-Fos quantification, the brain was imaged using a 30X magnification objective. Image stacks were individually analyzed, and c-Fos in the telencephalon was quantified using ImageJ Fiji software.^30^ A predefined area within the telencephalon was selected and consistently utilized for the analysis across all images. This procedure was conducted twice, and the statistical analysis was performed on the pooled data using GraphPad Prism 10.2.1 software (GraphPad Software, CA, USA).

### Mendelian randomization

#### Datasets

We used hypertension summary statistics with 1,237 cases and 359,957 controls provided by FinnGen^31^ and a recent meta-analysis comprising 38,691 individuals with ADHD and 186,843 controls^32^ as outcomes.

The manually curated list of traits includes blood cell counts, metabolites, amino acids, lipids, and enzymes. A total of 140 GWAS summary statistics were used as exposures for the MR experiments, covering measurements of the body, enzymes, blood cells and their proteins, lipids, fatty acids, as well as small molecules, such as amino acids, ketones, and glycolysis metabolites (Supplementary Table 1). All exposure datasets, with the exception of estimated glomerular filtration rate (eGFR) summary statistics, were generated using samples from individuals of European ancestry in the UK Biobank.^33^ Of the 140 exposure datasets, 78 were produced by Nightingale Health in their disease-wide association scan of plasma nuclear magnetic resonance (NMR) biomarkers for 118,444 individuals.^34^ An additional 45 datasets were produced by The Neale Lab (http://www.nealelab.is/uk-biobank/) following their GWAS analysis (version 3) of over 7,000 phenotypes using samples from 361,194 individuals. For three traits - neutrophil count over lymphocyte count, platelet count over lymphocyte count, and aspartate aminotransferase over alanine aminotransferase - GWAS were run in-house based on UK Biobank data. The dataset for estimated Glomerular Filtration Rate (eGFR) was produced by Wuttke *et al.*^35^

#### Data pre-processing

The datasets were pre-processed with version 1.7.8 of the tool MungeSumstats,^36^ using default parameters. These include checking to ensure variants are on the respective human reference (GRCh38/Hg18 or GRCh37/Hg19 depending on the datasets), removing duplicate SNPs, and removing multi-allelic SNPs (according to the SNP database db144).

#### Mendelian randomization

First, pre-processed summary statistics files were read and thresholds were applied to the exposure SNPs to ensure they were valid instruments. The *p*-value threshold was *p* < 5E−08 for genome-wide experiments and *p* < 1E−04 for drug-target experiments. Independent SNPs were identified by performing linkage disequilibrium (LD) clumping to discard SNPs in LD with another variant with a smaller *p*-value association based on the European reference panel from the 1000 Genomes Project (https://www.internationalgenome.org/home) using the ld_clump() function of the R package ieugwasr (https://github.com/MRCIEU/ieugwasr), which provides a wrapper around PLINK.^37^ Variants were only included that are correlated at r^2^ < 0.001 for genome-wide, or r^2^ < 0.1 for drug-target MR experiments. The window was set to 10Mbp in both cases, which is the default used by the ld_clump() function. If at least three SNPs remained as instrumental variables, the MR-Rücker framework^38^ was utilized to decide whether IVW or MR-Egger is best supported by the data. Otherwise, the Inverse-variance weighted (IVW method) for two SNPs or the Wald ratio method (one SNP) were used.^39^ All methods were implemented in the TwoSampleMR package.^40^

In order to take the multiple drug targets of amlodipine into consideration, we extended the common practice in drug-target MR to draw variants from genes that are associated with an exposure by providing multiple gene ranges from which variants could be selected. We provided flanks of 5 kbp around each gene large enough to try and capture as many relevant variants, but with as little overlap with neighboring genes as is feasible.

Effect estimates correspond to the log odds of the outcome per copy of the effect allele.

### Polygenic Risk Score Analysis

The UK Biobank is a prospective study of over 500,000 UK residents recruited from 2006 to 2010. Upon initial assessment, participants responded to socio-demographic, lifestyle, and health-related questions via a touch-screen questionnaire and an interview before completing a range of physical measures and providing blood, urine, and saliva samples, which were the basis for biomarker assays and for genotyping and imputation. Longitudinal data consists primarily of linkages with national datasets including hospital inpatient health records.

Starting from summary statistics for an ADHD GWAS of a European population^41^ and filtering for genetic variants available in the UK Biobank imputation data, we applied the PRS-CS-auto algorithm to infer posterior effect estimates for these variants using an external linkage disequilibrium reference panel for a European population.^42^ These estimates, along with the individual-level UK Biobank imputation data, were the basis for the computation of individual-level, per-chromosome ADHD polygenic risk scores for UK Biobank participants using PLINK 2.0^43^. Individual-level genome-wide risk scores were then computed as the weighted mean of per-chromosome scores, with weights calculated at the individual-level to account for within-chromosome missing variant data. That is, 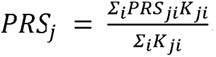, where *PRS_ji_* and *K_ji_* represent, respectively, for an individual *j*, the mean posterior effect estimate for chromosome *i* and the number of variants present in *i*. Risk scores were subsequently standardized for convenience.

Prior to analysis, the UK Biobank cohort was subject to quality control filters to exclude individuals missing data for sex (as determined from genotyping analysis) or assessment age, cases of sex chromosome aneuploidy, outliers for heterozygosity or missing rate, and individuals of non-European ethnic background. We identified case status for medications and health outcomes of interest on the basis of self-reports at assessment. Alternative indicators for health outcomes were identified on the basis of ICD10 codes registered in the longitudinal hospital inpatient data. Finally, case status for mood swings and risk-taking were identified on the basis of responses, respectively, to touchscreen questions “Does your mood often go up and down?” and “Would you describe yourself as someone who takes risks?”

To assess associations of ADHD risk scores with health outcomes of interest, we trained multiple logistic regression models on our UK Biobank cohort with ADHD risk score as an independent predictor. To assess the effects of amlodipine on mood swings and risk-taking behavior, we trained a logistic regression model for each outcome with ADHD risk score and amlodipine status as independent predictors.

## Data Availability

The data that support the findings of this study are available from the corresponding author upon reasonable request.

## Acknowledgement

3Z is supported by Icelandic Technology Development Fund grant no. 2220781-601. The research work at biotx.ai GmbH was supported by the Investitionsbank des Landes Brandenburg (ILB), the European Regional Development Fund (ERDF), and the European Social Fund+ (ESF+). The research has been conducted using the UK Biobank Resource under Application no. 36226.

## Author contribution

D.Þ.H., H.S.S. ran larval assays; M.P., D.Þ.H. ran blood-brain barrier penetration assay; B.G. ran c-Fos study; C.H., J.R.-T. ran 5-CSRTT assay; D.Þ.H., H.S.S., B.G., and H.Þ. analyzed data; H.A.B., M.F.S. ran MR analysis; J.L.C., M.F.S. ran PRS analysis. K.Æ.K. conceived experiments. M.P., H.Þ., M.F.S., H.A.B. and K.Æ.K. designed experiments. All authors edited the manuscript; K.Æ.K. wrote the manuscript.

