## Supplementary figures and images for "Validation of L-Type Calcium Channel Blocker Amlodipine as a Novel ADHD Treatment through Cross-Species Analysis, Drug-Target Mendelian Randomization, and clinical evidence from medical records"

### Supplementary Figure 1

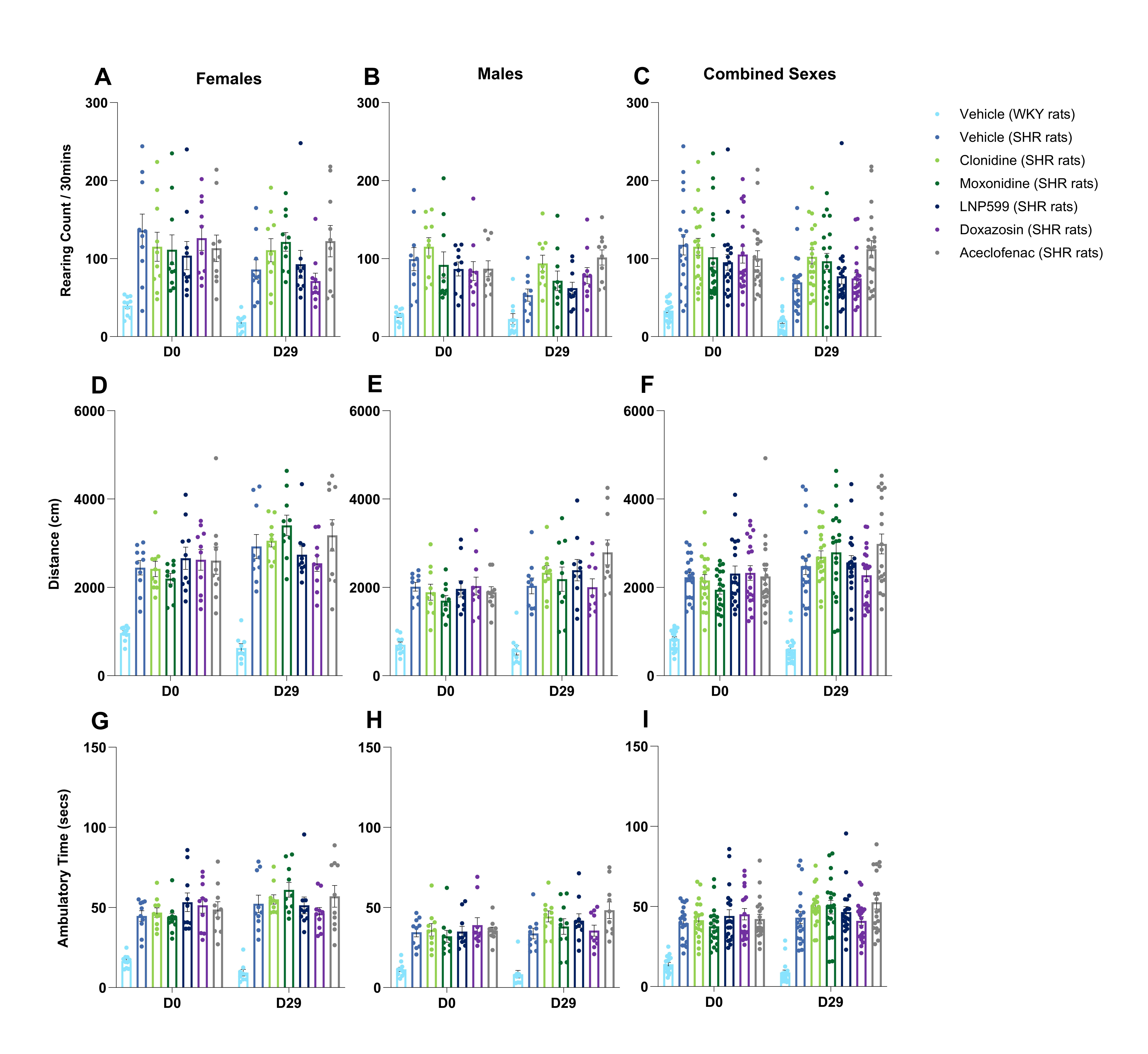

### Supplementary Figure 2

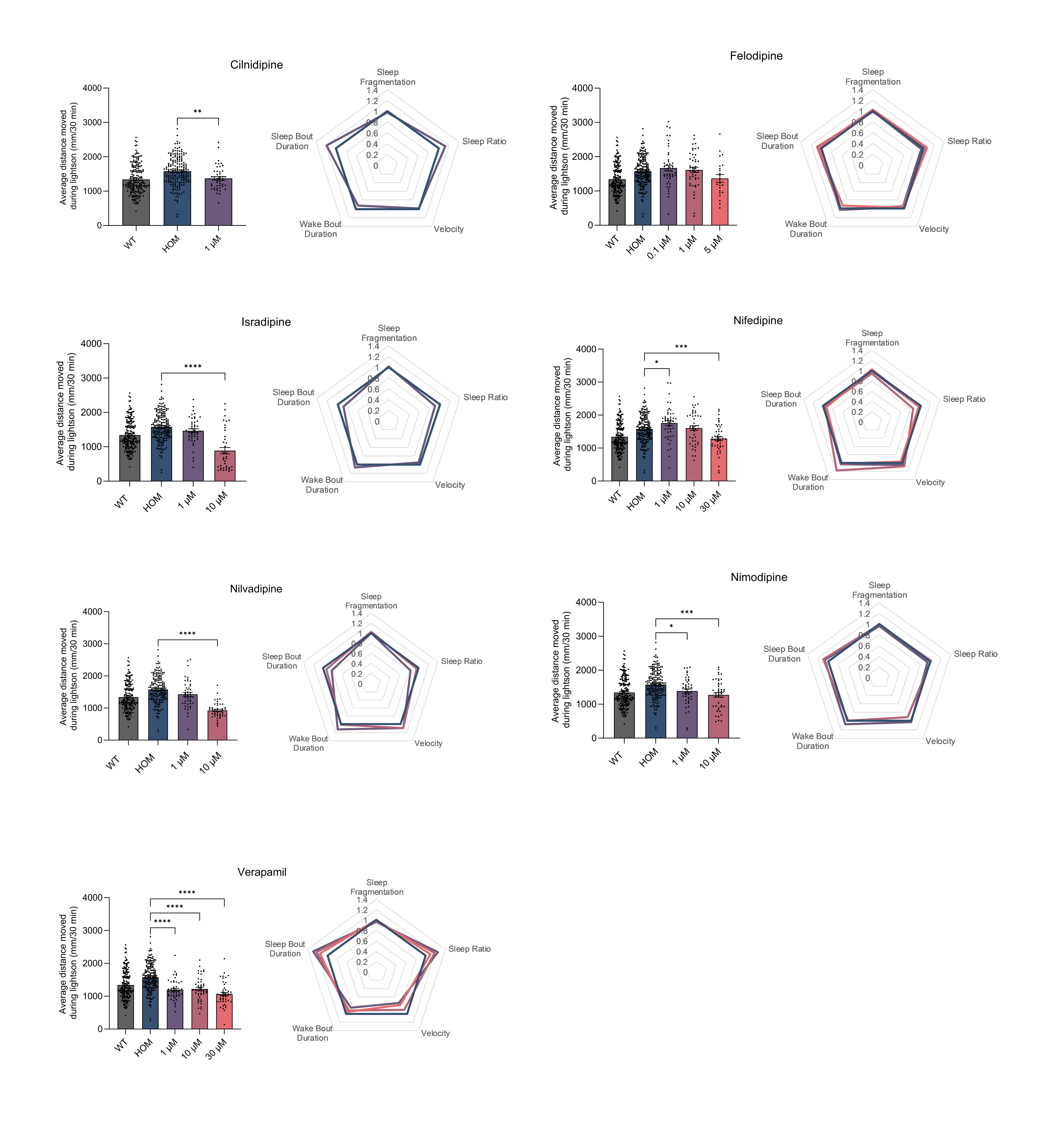

### Supplementary Figure 3

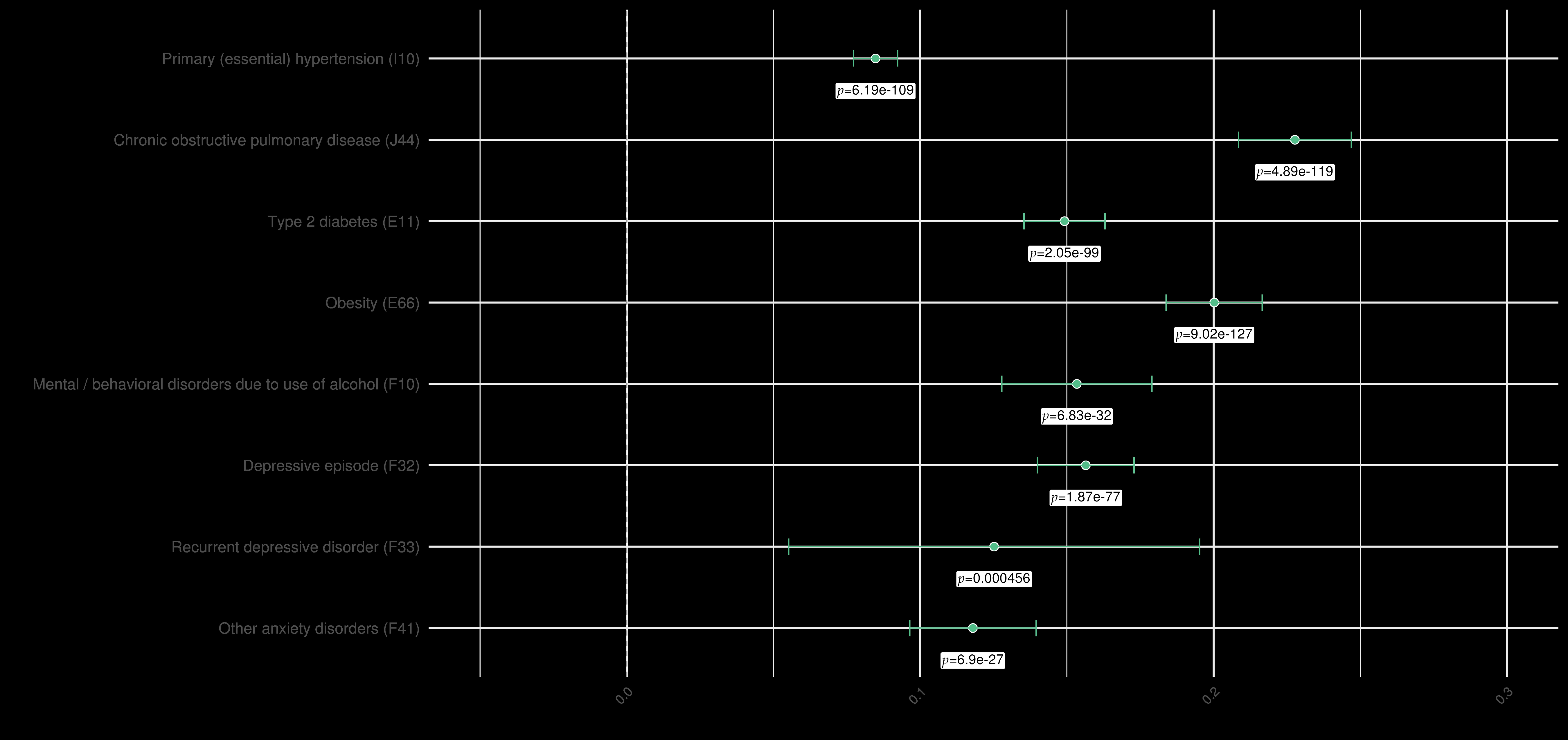

### Supplementary Figure 4

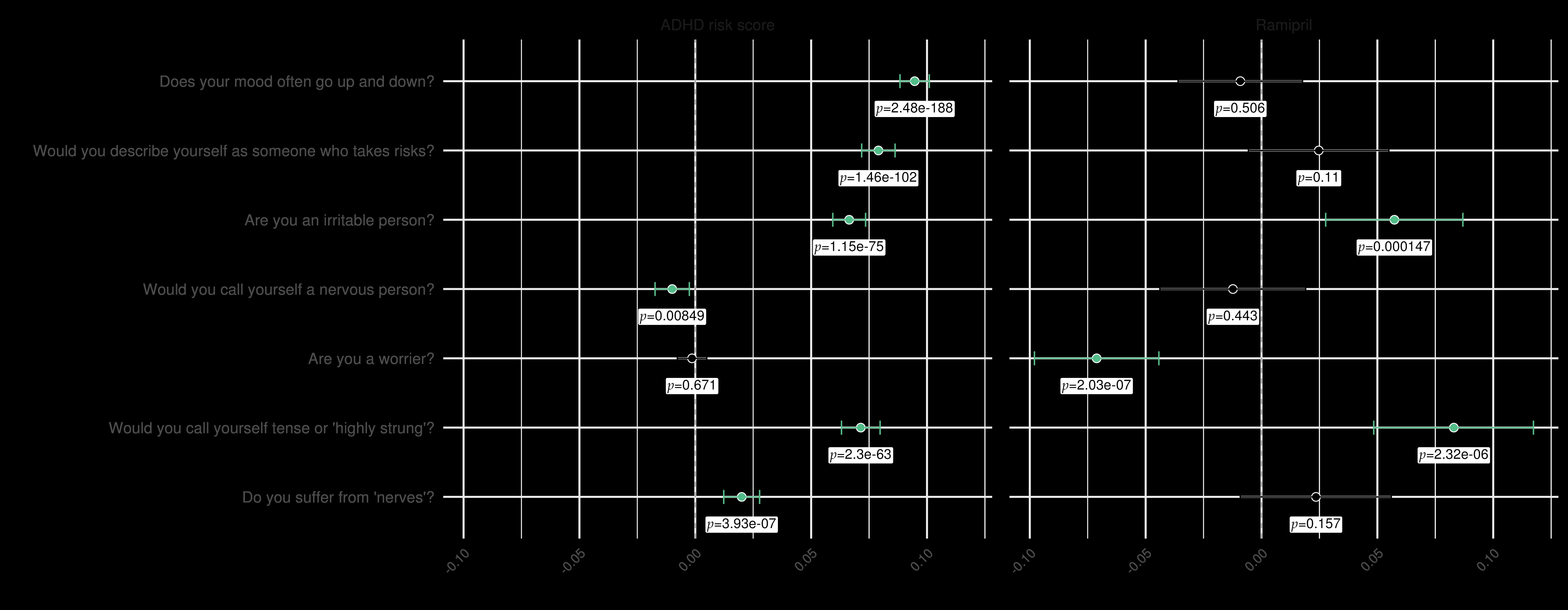
